# Knowledge, Attitude, and Practice among school students and teachers about the Human Papillomavirus vaccine in Central Kerala- a cross-sectional study

**DOI:** 10.1101/2025.02.24.25322810

**Authors:** Thomas Pious, Anjum John, V.R Reshma, Mercy John Idicula

**Author notes:** Corresponding Author: Anjum John.

## Abstract

**Background:** Human Papillomavirus (HPV) is the fourth most common cancer among women globally and a major public health concern. Uptake of HPV vaccine remains low despite its availability, in several countries, including India. India planned to introduce the HPV vaccine into the Universal Immunization Program (UIP), aiming to improve vaccine coverage. Knowledge, attitude, and practice (KAP) regarding the HPV vaccine among school-going adolescents, their parents, and teachers remain largely unexplored in Kerala, despite its high literacy. Understanding the awareness and perceptions of these stakeholders is critical for the successful implementation of vaccination programs.

**Methods:** This cross-sectional study will assess the KAP regarding HPV vaccination among school students (aged 11–15 years), their parents, and teachers in Thiruvalla Municipality, Kerala. A structured, validated questionnaire will be used to collect data from randomly selected government, private, and aided schools. Stratified random sampling will be used for student selection, while all teachers in the selected schools will be included through census sampling. Parents will be recruited using a convenience sampling approach. Descriptive and inferential statistical analyses will examine KAP levels, compare demographic groups, and identify predictors of vaccine acceptance.

The study has received ethical approval and currently is in the process of obtaining necessary permissions from school authorities and fulfilling administrative requirements for implementation.

**Expected Results:** We anticipate finding gaps in knowledge and varying levels of acceptance of the HPV vaccine among students, parents, and teachers. While some stakeholders may be aware of HPV and its link to cervical cancer, misconceptions and cultural barriers may hinder vaccine uptake. We expect that health education with access to reliable information will be associated with higher knowledge and a positive attitude toward vaccination.

**Expected Conclusions:** The study will provide insights into existing knowledge gaps and barriers to HPV vaccination in Kerala. Findings will inform the design of targeted educational interventions and policy recommendations to improve vaccine acceptance and coverage. Strengthening school-based awareness programs and involving teachers and parents as key advocates may significantly enhance HPV vaccine uptake and contribute to cervical cancer prevention efforts in India.

## BACKGROUND

The Human Papilloma Virus (HPV) is a group of 200 known viruses that are found in sexually active men and women. It is the most common sexually transmitted virus around the world. Most of these viruses are naturally managed by the human immune system and pose no significant concern.(1) About 40 types of HPV virus strains affect the anogenital region. Many of the HPV infections are cleared by the immune system without complications. Some high-risk strains are associated with cancers such as cervical, anal, oropharyngeal, and penile cancer. Despite the availability of effective vaccines, HPV-related diseases remain a significant public health concern, particularly in low- and middle-income countries where screening and vaccination coverage may be low. Understanding public knowledge, attitudes, and practices regarding HPV and its vaccine is crucial in developing effective prevention strategies. (2–5)

### Public health problems due to the HPV virus

Cervical Cancer is a major public health problem; on a global scale, being the fourth most common cancer among women. In 2023, it was estimated that there were 604,000 new cervical cancer cases, causing around 342,000 deaths worldwide. In the 12-year period between 2018-2030, the number of cervical cancer cases has been projected to increase by about 100,000 cases each year. Illiteracy and early sexual activity are risk factors for more than 85% of those affected by cervical cancer.(6) The survival and well-being of children are at risk when their mothers die from cervical cancer.(7)

### Disparities in health care around the world

Cervical cancer is a preventable and curable disease. Despite the availability of the HPV vaccine, - a key tool for the prevention of cervical cancer caused by HPV, and effective treatment measures-much of the world cannot avail of these services. In some regions with the highest disease burden of cervical cancer, there has been poor implementation of prevention and treatment measures. Statistics show that deaths due to cervical cancer are three times higher in low- or lower-middle-income countries. Cervical cancer is a disease that demonstrates the widespread disparities in the availability, accessibility, and adaptability of health care. (8)

HPV vaccination, cervical cancer screening, and management of cervical cancer must be integrated in strategic ways into the health care system by involving people in their own health care. (9)Till such time, it will continue to wreak havoc as one of the most common cancers and a major cause of cancer death in women. (10)

### History of the HPV vaccine

The first HPV vaccine was licensed for use in 2007 in a few countries of the world. Despite initial skepticism about a vaccine for a sexually transmitted disease, slowly but surely, vaccine uptake has increased. Prevention of HPV would, therefore, reduce the incidence, morbidity, mortality, and costs associated with cervical cancers as well as genital warts. Screening and treatment of precancerous lesions, and prophylactic vaccination against HPV can all work together in effectively reducing the burden of such diseases in cost effective ways. (11)The success of these vaccines in preventing cervical cancer has led to a global strategy that aims to eradicate cervical cancer.

### Global strategy for the elimination of cervical cancer

Public health microbiology is at the threshold of a rare but doable feat-the elimination of a non-communicable disease-cervical cancer. Through the Global Strategy to eliminate cervical cancer adopted in 2020, there is a strategic plan to eliminate cervical cancer as a public health problem by 2030. Countries have been given the 90-70-90 target strategy towards elimination of cervical cancer. This strategy aims to vaccinate 90% of girls less than 15 years of age with the HPV vaccine. By 35 years of age and again by 45 years of age, 70% of women must be screened with a high-performance test to detect cervical cancer. Further, 90% of women identified with cervical disease should receive treatment. (9)

### HPV virus in India

India is a lower middle-income country. The vastness of the country and the diversity of its population are responsible for the lack of proper documentation of the epidemiology and patterns of HPV virus strains. Thus, as in other lower income countries, the implementation of prevention and control programs has been difficult. In India, annually, about 1,32,000 new cervical cancer cases and 80,000 deaths occur, and the prevalence of HPV type 16 was found to be extremely high.(12) At present India has the highest burden of cervical cancer in the world. (13)The importance of the HPV vaccination cannot be overstated in this context.

### Vaccine coverage in India

Since 80% of cancers caused by HPV can be effectively prevented by the vaccine, it offers a window of opportunity for controlling this disease. India has two HPV vaccines licensed for use since 2008.(13,14) Both the quadrivalent and the bivalent vaccines offer considerable protection against HPV induced cancer and for a long duration (about 10 years).

The vaccine was first introduced in the State of Sikkim, in 2018 as part of a state-wide school-based immunization program targeting adolescent girls. The initiative achieved high vaccination coverage due to strong governmental support, school-based delivery, effective collaboration between health and education departments, and robust social mobilization strategies, and effective awareness campaigns. The state achieved over 95% vaccination coverage among targeted girls for both doses through two campaigns, with no severe adverse events reported. The vaccine was well accepted by all stakeholders, with minimal refusal observed. (15)

The indigenously developed quadrivalent vaccine was recommended for use in the UIP as a two-dose regimen for girls. The National Technical Advisory Committee on Immunization in 2022(16) recommended the introduction of the HPV vaccine as part of the Universal Immunization (UIP) program with the first dose at 9 years of age and a onetime catch-up immunization at 9-14 years for girls.

Boys will be immunized once 80% coverage for girls is achieved. The vaccine was planned to be rolled out for use free of charge in the second half of 2024. (17) And, two districts of Kerala, Wayanad and Alappuzha were planned to be included in the pilot phase of the launch of this vaccine. (18)Though the nationwide implementation of the HPV vaccine in India under the UIP is facing technical delays, efforts are on.(19)

### Vaccine coverage in Kerala

The exact statistics on the voluntary use of the HPV vaccine in Kerala are not available.

The prevalence of cervical cancer has shown a declining trend according to some studies with rates as low as 7-9 cases per 100, 000 per year. This is used sometimes as an argument against the introduction of the HPV vaccine in the state and to increase the use of cervical cancer screening techniques rather than vaccination.(20) One study has shown an 8% coverage of medical students in Kerala in 2018.

The *Happy Noolpuzha Initiative*, launched by Noolpuzha Gram Panchayat in Wayanad in March 2024, provided free HPV vaccination to girls aged nine and above, making it the first such program in a tribal panchayat in India. With an initial vaccination of 53 girls and plans to expand, the initiative aims to combat cervical cancer by ensuring widespread immunization, highlighting the community’s proactive commitment to women’s health.(21)

A study by Backer, Sudha et al published in 2024 conducted in about 1000 college students from Kollam district of Kerala(22) showed that only 48.5% of students had heard about the HPV vaccine.

The knowledge and attitude towards acceptance of these vaccines in India is not well studied. (10,23)

### Research Hypothesis

The hypothesis of this research is that knowledge, awareness, and uptake of the HPV vaccine among school-going children, parents, and teachers in Kerala are low. There is a significant need for targeted educational interventions, and community-based initiatives to improve knowledge and acceptance among the vaccine-target age group.

### Research Question

This study seeks to answer questions about what the level of knowledge is, attitudes and practices related to HPV vaccination are, among key stakeholders (students, teachers, and parents) in Thiruvalla Municipality.

### Aim of the research

The study seeks to evaluate the knowledge, attitudes, and current practices related to the HPV vaccine among school-going children, their parents, and teachers in Thiruvalla Municipality. Teachers, parents, and the adolescent school going children are the stakeholders in the HPV vaccination program.

### Rationale

There is limited information on the knowledge, attitudes, or the current practices related to the vaccine among the adolescent school-going children and their teachers. A review of existing literature revealed a lack of data on HPV vaccines related knowledge, attitudes, and practices in schools. Understanding the knowledge, attitudes, and practices of HPV vaccination among school children is crucial for several reasons. It helps in assessing the effectiveness of vaccination programs, identifying potential barriers to vaccination acceptance, and informs the development of targeted educational interventions to enhance uptake of the vaccines.

### Relevance

Cervical cancer is a significant global health concern. As part of the WHO Global Strategy to eliminate cervical cancer as a public health problem, achieving adequate knowledge, fostering a positive attitude, and ensuring that at least 90% of girls receive the HPV vaccine by 15 years of age are critical goals.

The Sustainable Development Goals(SDGs) advocate that no one be left behind, and the successful implementation of this elimination goal can prevent 74 million new cases of cervical cancer and avert 62 million deaths from cervical cancer, it is essential that the beneficiaries of the vaccine be aware of the virus, the disease and its vaccine.(24) To achieve this, it is essential that vaccine target groups are well-informed about HPV, cervical cancer, and available preventive measures like vaccines.

Assessing the knowledge, attitude, and practice among school children plays a vital role in public health by promoting early awareness, fostering a positive attitude toward HPV vaccination, and reducing the risk of HPV-related diseases in adulthood. The findings of this study can inform policymakers, by helping them take evidence-based decisions to make policy decisions, to strengthen HPV vaccination programs and improve public health outcomes.

## OBJECTIVES

The study aims to assess the knowledge, attitude, and practice on HPV vaccination among school children (aged 11-15 years), their teachers, and parents, of Thiruvalla Municipality. The study will use the findings to design targeted health education programs, addressing the specific needs of school-going students, their teachers, and parents in Thiruvalla Municipality.

### Review of literature

Several studies highlight the low levels of awareness and uptake of HPV vaccination in India. A study conducted among college students in Kollam, Kerala, found that while 85.6% of students were aware that HPV causes cervical cancer, only 48.6% had heard of the HPV vaccine, and vaccination rates remained low (11.6% in medical students, 4.9% in non-medical students) (22) Similarly, research by Basu et al. (2021) among adolescent girls in North India found that vaccine uptake was hindered by lack of knowledge, financial constraints, and cultural hesitancy.(25).

HPV vaccination programs in schools have been identified as an effective way to increase vaccine awareness and acceptance (26) However, studies indicate that many school-going children lack adequate knowledge about HPV and its vaccine. Research from Maharashtra revealed that only 2.5% of school students had heard of the vaccine, and this improved after an awareness class was conducted.(27) Teachers play a crucial role in disseminating vaccine-related information, yet studies suggest that many lack adequate training on HPV and its prevention.(28) Parental attitudes also significantly influence vaccination rates, with concerns over vaccine safety and cultural stigma acting as major barriers.(29)

Sikkim was the first Indian state to introduce the HPV vaccine in 2018, achieving over 95% vaccination coverage through a school-based program supported by the government and healthcare professionals.(15)More recently, the Happy Noolpuzha Initiative in Wayanad (2024) has set a precedent by providing free HPV vaccination in a tribal panchayat, demonstrating how community-led efforts can improve vaccine acceptance.(30)

Studies emphasize that improving vaccine uptake requires comprehensive health education initiatives targeting students, teachers, and parents. (31) Health campaigns incorporating school-based awareness programs, parental counseling, and teacher training have been found effective in improving HPV vaccination rates.(32)

Given the gaps in knowledge and vaccine uptake in Kerala, a targeted educational intervention could significantly enhance awareness and acceptance of HPV vaccination.

## Methods

### Study design

This study will employ a cross-sectional study design to assess the knowledge, attitude, and practices (KAP) related to HPV vaccination among school-going children (aged 11-15 years), their teachers, and parents in Thiruvalla Municipality. A cross-sectional study is appropriate for this research as it allows for the collection of data at a single point in time, providing insights into the current state of HPV vaccine awareness and uptake among different target groups. It is cost-effective and requires fewer resources compared to longitudinal studies while still providing valuable data for planning health interventions.

### Study setting

The study will be conducted in higher secondary schools within Thiruvalla Municipality, specifically targeting students aged 11-15 years, their teachers, and parents. Schools will be randomly selected from different administrative zones of the municipality to ensure representation of various demographic groups.

Thiruvalla is a prominent municipality in Pathanamthitta district, Kerala, India, known for its high literacy rates, well-established educational institutions, and strong healthcare infrastructure. The municipality is home to several higher secondary schools that cater to students from diverse socio-economic and cultural backgrounds. These schools are affiliated with different educational boards, including the Kerala State Education Board, CBSE (Central Board of Secondary Education), and ICSE (Indian Certificate of Secondary Education).

The sample frame will include:

- Government and government-aided schools.
- Private and unaided schools.
- Schools affiliated with different educational boards (State Board, CBSE, ICSE).

A mix of urban and semi-urban schools will be included to capture variations in awareness, attitudes, and practices regarding HPV vaccination.

### Study period

The study will be conducted over a 12-month period following the receipt of institutional ethical clearance, which has been obtained. The timeline will be structured to allow for the phased implementation of the study, ensuring that all necessary processes are conducted systematically and within a reasonable time.

### Study participants

#### Inclusion Criteria

The study will include:

- All school students aged 11 years and above are enrolled in the higher secondary schools of Thiruvalla Municipality and consent to participate in the study.
- All teachers who teach students aged 11 years and above are willing to participate in the study.

#### Exclusion Criteria

The study will exclude:

- Students absent on the day of data collection: Any student who is not present during the data collection period will not be included in the study.
- Teachers who are absent on the day of data collection: Teachers who are not available during the data collection period will be excluded.

A total of 4 schools were selected from the list of schools in Thiruvalla Municipality provided above. The schools are chosen using a random selection method from different educational categories: government, government-aided, and private schools, ensuring representation across different types of institutions. The schools selected will be as follows:

- BL School
- BC Res School
- MT Res School
- C Cen School

Stratified Random Sampling will be used to select students from the chosen schools.

- The schoolchildren will be grouped based on their age (11-15 years).
- A proportional sample from each age group (11–12, 13–14, 15) will be drawn to ensure that each subgroup is well represented in the study.
- From each class in the selected schools, students will be randomly selected for participation. This helps ensure that the sample includes students from different backgrounds, school subjects, and educational boards. Until the sample size is reached.

A census sampling approach will be used for teachers in the selected schools. All teachers who are responsible for students in the 11–15-year age group will be included in the study, ensuring that teachers with direct interaction with the target group are represented. Teachers will be invited to participate voluntarily, and a complete enumeration will be conducted for this group.

Sampling Method for Parents: For the parents of students aged 11-15 years, convenience sampling will be used. The study will engage parents during parent-teacher meetings or through communication channels established by the school (e.g., WhatsApp groups, Google Forms). A sample of parents/guardians will be selected based on their availability and willingness to participate in the study.

### Sample size

Sample size was calculated using the proportion of students with awareness of HPV vaccine (p=18.3%) from previous study (Fagbule et al, 2020), confidence level (1-α) as 95%, absolute precision (d) as 5%. The sample size obtained was 230 using the formula:

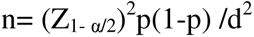

This sample size was multiplied with a design effect (DEFF) of 1.5. Thus, the final sample size is **345** (i.e., 230*DEFF).

### Data Collection plan

A structured questionnaire has been developed and validated for reliability through a pilot study prior to its use in the main study. The next step will involve contacting the school authorities of the selected schools for permission to conduct the survey.

- For government schools, permission will be sought from the District Chief Education Officer (DCDS).
- For private schools, consent will be obtained from the respective school authorities.

Once permission is granted, the study will be explained to the school authorities, and their formal consent will be obtained. Since the students are minors, written consent from parents or guardians will also be required for student participation.

The questionnaire will be administered in English or Malayalam, based on the language preference of the students, to ensure that they can understand the questions fully. This will allow for more accurate and reliable responses.

The teachers of the students in the selected schools will also be invited to participate in the study. With their consent, the same questionnaire will be administered to the teachers.

Data will be collected using paper and pen forms, ensuring confidentiality throughout the process. The information will then be entered into a Google Sheet or Excel spreadsheet for data analysis. These files will be password-protected, and only authorized study investigators will have access to the data.

### Ethical considerations

The virus and vaccine related questions can be of a sensitive nature and potentially embarrassing for students in the adolescent age group to answer. Special care will be taken to address any concerns. Since it is important to obtain this information, parental permission and consent will be obtained to ensure adequate protection for minors involved as study participants. Study investigators will be on hand to answer questions that may come up during the study. This will help ensure that participants feel comfortable and informed throughout their involvement.

Data will be stored in password protected files and the privacy of the participants will be protected to the extent possible. Only authorized study investigators will have access to the data. Every effort will be made to ensure the privacy of the participants is maintained throughout the study, and personal information will not be disclosed without explicit consent.

### Data Analysis

The data analysis for this study will be performed in a systematic and rigorous manner, ensuring that the findings are reliable. The analysis will address the research objectives related to the knowledge, attitudes, and practices (KAP) regarding HPV vaccination among school children, teachers, and parents in Thiruvalla Municipality. The following steps outline the planned data analysis process:

1. Data Cleaning and Preparation

- Data Entry: The data collected through the structured questionnaires will be entered into a Google Sheet or Excel spreadsheet. All personal identifiers will be removed to ensure confidentiality.
- Validation and Cleaning: The data will be thoroughly checked for completeness, accuracy, and consistency. Any incomplete or erroneous data will be flagged and cleaned by reviewing the original responses.
- Managing Missing Data: Missing data will be managed using appropriate methods, such as imputation or exclusion based on the extent of missingness (e.g., excluding cases with more than 20% missing data).
2. Descriptive Analysis

- Demographics: Descriptive statistics (frequencies, percentages, means, and standard deviations) will be used to summarize demographic variables, including:

○ Age
○ Gender
○ Socioeconomic status (if available)
○ School type (private vs. government)
- Knowledge: The responses regarding knowledge about HPV and the HPV vaccine will be summarized using:

○ Percentages of participants who correctly identify HPV as the cause of cervical cancer.
○ Percentages of participants who know about the vaccine’s target age group, schedule, and benefits.
- Attitude: Descriptive statistics will be used to summarize attitudes toward the HPV vaccine, such as:

○ Willingness to get vaccinated.
○ Willingness to recommend the vaccine to others.
○ Perceived barriers to vaccination.
- Practice: Data regarding vaccination practices will be summarized, including:

○ Percentage of participants who have been vaccinated.
○ Reasons for not getting vaccinated (e.g., lack of knowledge, cultural beliefs, cost).
3. Comparative Analysis

- Comparison of Knowledge, Attitude, and Practice (KAP) between Groups:

○ By Gender: t-tests or chi-square tests will be used to compare knowledge, attitudes, and practices between male and female students, teachers, and parents.
○ By Age Group: Analysis will be performed to compare KAP across different age subgroups within the 11-15 years group.
○ By School Type (Private vs. Government): t-tests or chi-square tests will be used to compare knowledge, attitude, and practice between students, teachers, and parents from private and government schools.
- Knowledge, Attitude, and Practice will also be compared among:

○ Schoolchildren vs. teachers vs. parents.
○ Participants who have had prior health education vs. those who have not.
4. Multivariable Analysis

- Logistic Regression: To identify predictors of HPV vaccination knowledge, attitude, and practice, multivariable logistic regression analysis will be performed. The following factors will be considered as potential predictors:

○ Socioeconomic status of students.
○ Gender.
○ Prior exposure to health education on HPV and cervical cancer.
○ Parental influence on vaccination decisions.
○ Teacher’s knowledge and attitude toward the HPV vaccine.
- Effect Modifiers and Confounders: Analysis will adjust for potential confounders (e.g., age, gender, socioeconomic status) and effect modifiers (e.g., school type, previous exposure to health education) to assess their influence on the outcomes.
5. Interpretation of Knowledge Scores

- Knowledge Assessment: A knowledge score will be calculated based on correct responses to key questions regarding HPV and its vaccine. The total score will be divided into categories such as:

○ Low knowledge
○ Moderate knowledge
○ High knowledge
- These categories will then be analyzed to identify factors associated with higher or lower levels of knowledge, using chi-square tests or logistic regression as appropriate.
6. Data Presentation

- Tables and Figures: Data will be presented using tables and bar graphs, with clear labels for each variable. The descriptive statistics for knowledge, attitudes, and practices will be displayed to give a comprehensive overview of the data.
- Statistical Significance: A p-value of <0.05 will be considered statistically significant for all hypothesis tests.
7. Sensitivity Analysis

- Missing Data: Sensitivity analysis will be performed to assess the impact of missing data on the results. This will involve comparing the findings with different methods of handling missing data, such as imputation or exclusion.

### Outcome measurements

The knowledge, attitude, and vaccine practices related to the HPV vaccine will be obtained from both teachers and students. The primary outcome measures for this study will assess the knowledge, attitude, and practice (KAP) related to HPV vaccination among the selected participants (school children, teachers, and parents) in Thiruvalla Municipality. The following specific outcome measures will be evaluated:

1. Knowledge:

○ Awareness about HPV and its association with cervical cancer.
○ Knowledge of the availability and benefits of the HPV vaccine.
○ Understanding of the target age group and vaccination schedule.
2. Attitude:

○ Willingness to get vaccinated or recommend the vaccine.
○ Perceptions of the vaccine’s safety and efficacy.
○ Cultural, social, or personal beliefs regarding vaccination.
3. Practice:

○ Uptake of the HPV vaccine (for students, teachers, and parents).
○ Barriers to vaccine uptake (e.g., cost, accessibility, stigma).
○ Sources of information about the vaccine (e.g., school programs, health workers, media).

Confounders: Variables that may influence the relationship between independent and dependent variables could include:

1. Socioeconomic Status: Students from different socioeconomic backgrounds may have varied access to healthcare and educational resources, affecting their knowledge and attitude toward vaccination.
2. Gender: Gender differences might influence attitudes and practices related to HPV vaccination, as some cultural factors may affect vaccine uptake among boys and girls differently.
3. Previous Exposure to Health Education: Prior educational interventions or awareness programs may influence knowledge levels and attitudes toward the HPV vaccine.
4. Parental Influence: Parental approval or opposition to vaccination could significantly influence children’s attitudes and practices regarding HPV vaccination.

Effect Modifiers: Variables that may alter the strength or direction of the relationship between the independent and dependent variables could include:

1. Age: Different age groups within the 11-15 years age range may have varying levels of understanding and acceptance of the HPV vaccine, which could modify the observed outcomes.
2. Type of School (Private vs. Government): The school type (private vs. government) may influence the level of access to health education resources and the degree of parental involvement, which could modify the knowledge, attitudes, and practices related to HPV vaccination.
3. Teacher Influence: Teachers’ attitudes toward HPV vaccination may modify students’ perceptions and behaviors, as teachers play an important role in influencing students’ health decisions.
4. Cultural and Religious Beliefs: Cultural or religious beliefs may modify how the HPV vaccine is perceived and whether it is accepted, potentially influencing the willingness to vaccinate.

Predictors: Variables that are expected to influence or predict the outcome measures include:

1. Educational Level of Parents: Parents with higher levels of education may be more knowledgeable and open to the HPV vaccine, influencing their children’s awareness and vaccine uptake.
2. Health Information Sources: The source of health information (e.g., schools, healthcare professionals, media) could predict the level of knowledge, attitudes, and practices toward the HPV vaccine.
3. Peer Influence: Peer recommendations or discussions about the HPV vaccine could predict students’ willingness to get vaccinated or recommend the vaccine to others.
4. Teacher Knowledge and Attitude: Teachers who have a good understanding of HPV and the vaccine are likely to influence students’ knowledge and attitudes toward vaccination, thus acting as predictors for students’ knowledge and practices.
5. Prior Vaccination Experience: Previous experiences with vaccinations (either personal or through family) may function as a predictor for HPV vaccine acceptance, as students and parents who have had positive experiences with vaccines may be more likely to adopt the HPV vaccine.

Reporting and Interpretation of Results

The findings will be summarized in a final report, which will include:

○ Descriptive statistics and comparisons between different demographic groups.
○ Identification of significant predictors and barriers to HPV vaccine knowledge, attitudes, and practices.
○ Key recommendations for health education programs targeting school children, teachers, and parents based on the results.

Operational Definitions

1. HPV: Human papilloma virus is a virus that belongs to the papilloma viridae family and is a double stranded DNA virus. About 30 of these HPV types are known to cause anogenital and oral infections. They can be of the high and low risk categories of viruses.
2. Cervical Cancer: is a cancer of the cervix caused by a high-risk category of the HPV virus.
3. HPV vaccine: The HPV vaccine is a recombinant vaccine, and two types are licensed for use in India, with a third indigenously produced one, soon to be available in the vaccination program.
4. HPV vaccination: The vaccine is given intramuscularly and the dosage for children less than 15 years of age is two intramuscular doses at an interval of six months. Those who are above 15 years of age and the immunocompromised are recommended 3 doses of the vaccine, over 6 months.

## Funding

Though the proposal was sent to the Indian Council of Medical Research for funding, the proposal was not funded. The proposal has no funding now. Efforts are on to find institutional seed funding for the proposal. If this is successful, the proposal will be sent again to the premier funding agency with the preliminary results of the study.

Timeline

Gantt Chart for HPV Vaccination Study

| Task | Month<br>1 | Month<br>2 | Month<br>3 | Month<br>4 | Month 5 | Months 6-12 |
| --- | --- | --- | --- | --- | --- | --- |
| 1. Institutional Ethical Clearance | ✓ |  |  |  |  |  |
| 2. Pre-Study Preparations (1 month) | ● |  |  |  |  |  |
| Finalizing Study Tools | ● |  |  |  |  |  |
| Training of Data Collectors & Supervisors |  | ● |  |  |  |  |
| Seeking Permissions & Obtaining Consent |  | ● | ● |  |  |  |
| 3. Data Collection (3 months) |  |  |  | ● | ●<br>● |  |
| Student Surveys |  |  |  | ● | ● | ● |
| Teacher and Parent Surveys |  |  |  | ● | ● | ● |
| 4. Data Analysis (1 month) |  |  |  |  |  |  |
| Data Cleaning & Entry |  |  |  |  |  | x |
| Data Analysis |  |  |  |  |  | x |
| 5. Report Writing & Dissemination (1 month) |  |  |  |  |  | x |
| Preparation of Findings |  |  |  |  |  | x |
| Dissemination of Results |  |  |  |  |  | x |
| Legend: ✓ = Completed task ● = Ongoing tasks |  |  |  |  |  |  |

## Data Availability

All data produced in the present study will be made available upon reasonable request and will be available with the authors. As such it is a proposal yet.

## Declarations

Ethics approval and consent to participate: Ethical approval was sought and granted by the Institutional Review Board of the institution. (details will be provided on request). Information consent will be obtained from all participants before data collection. For minors, assents will be sought along with parental consent, and teachers will provide separate consent for their participation. Confidentiality and voluntary participation will be ensured throughout the study.

## Consent for publication

Will be taken from participants at the time of data collection.

- **Availability of data and materials**: All data generated or analyzed during this study are included in this published article and its supplementary information files. No publicly archived datasets will be analyzed or generated during the study.

## Competing interests: None that we can identify

- Funding: None, no institutional or agency support currently. Seed funding from the institution is sought.

## Authors’ contributions

TP is the student investigator on this proposal. He played a crucial role in the development of this research proposal. He contributed to the formulation of research objectives, review of relevant literature, and submission to the funding agency. Additionally, he was actively involved in drafting the informed consent process and forms, refining data collection tools, and ensuring ethical considerations were appropriately addressed.

MJ reviewed the proposal and provided constructive feedback at each stage of its development. She helped in refining the study design, ensuring methodological rigor, and addressing scientific and technical aspects to enhance the overall quality of the proposal.

AJ conceived the proposal and played a key role in its development, overseeing the writing process and ensuring adherence to timelines for funding submissions. She conducted a thorough literature review along with TP, provided critical feedback, and supervised the proposal’s refinement, ensuring its scientific rigor and overall quality before final submission.

RVR, the biostatistician is currently unattached to any institution and moved to the United Arab Emirates at this time-she contributed by critiquing the proposal, is expected to help with the data analysis and writing the results, reading the manuscript critically and providing general support for the project along with approval of the manuscript for final submission.

## Acknowledgements

None

## Animal or human data or tissue

“Not applicable”

## Student Questionnaire

The Human Papillomavirus (HPV) is a common virus that can cause certain types of cancers later in life. The HPV vaccine helps protect against infections that may lead to these diseases. Many countries recommend vaccination during adolescence for the best protection. This survey aims to understand what students know about the HPV vaccine. Your responses will help us find better ways to share important health information with young people like you. This is not a test—there are no right or wrong answers. Please answer honestly based on what you know.

For each question, please choose the answer that best matches what you think.

1. Does the HPV vaccine help protect girls from getting cervical cancer?

a. Yes
b. No
c. Maybe
d. I don’t know
2. Is it best to get the HPV vaccine between the ages of 9 and 26?

a. Yes
b. No
c. Maybe
d. I don’t know
3. Are different types of HPV vaccines available in our country?

a. Yes
b. No
c. Maybe
d. I don’t know
4. If a girl gets the HPV vaccine, does she still need to get tested for HPV later?

a. Yes
b. No
c. Maybe
d. I don’t know
5. Is the HPV vaccine more effective when given at a younger age?

a. Yes
b. No
c. Maybe
d. I don’t know
6. Is it recommended to take the HPV vaccine early for better protection?

a. Yes
b. No
c. Maybe
d. I don’t know
7. Can people still take the HPV vaccine if they haven’t taken it at a younger age?

a. Yes
b. No
c. Maybe
d. I don’t know
8. Does the HPV vaccine work the same way no matter what age someone gets it?

a. Yes
b. No
c. Maybe
d. I don’t know
9. Can the HPV vaccine help prevent HPV infections and reduce the risk of cervical cancer?

a. Yes
b. No
c. Maybe
d. I don’t know
10. Do people need to be tested for HPV before they get the vaccine?

a. Yes
b. No
c. Maybe
d. I don’t know
11. Is cervical cancer one of the most common cancers in women?

a. Yes
b. No
c. Maybe
d. I don’t know
12. Is HPV infection one of the main causes of cervical cancer?

a. Yes
b. No
c. Maybe
d. I don’t know
13. Can HPV be spread through close personal contact?

a. Yes
b. No
c. Maybe
d. I don’t know

Teacher Questionnaire on HPV and Vaccination

HPV is a widely prevalent virus, and some of its strains are linked to cervical and other cancers. The HPV vaccine is a preventive measure that has been introduced in many countries to reduce the burden of HPV-related diseases. As teachers, you play a key role in shaping students’ awareness of health-related topics. This survey seeks to understand your knowledge and attitudes about the HPV vaccine and its role in preventing HPV-related conditions. The findings will help improve health education strategies and ensure that students and parents receive accurate information about this important vaccine. Your responses will remain confidential.

For each question, please choose the answer that best matches your understanding.

1. Does the HPV vaccine provide protection against cervical cancer in women?

a. Yes
b. No
c. Maybe
d. I don’t know
2. Is HPV vaccination ideally recommended between the ages of 9 and 26?

a. Yes
b. No
c. Maybe
d. I don’t know
3. Are bivalent, quadrivalent, and Nona valent HPV vaccines available in our country?

a. Yes
b. No
c. Maybe
d. I don’t know
4. Should women who receive the HPV vaccine still undergo cervical cancer screening?

a. Yes
b. No
c. Maybe
d. I don’t know
5. Is the HPV vaccine most effective when given before exposure to the virus?

a. Yes
b. No
c. Maybe
d. I don’t know
6. Is it recommended to administer the HPV vaccine before the initiation of sexual activity to ensure maximum protection?

a. Yes
b. No
c. Maybe
d. I don’t know
7. Can individuals who are already sexually active still benefit from receiving the HPV vaccine?

a. Yes
b. No
c. Maybe
d. I don’t know
8. Does the effectiveness of the HPV vaccine decrease significantly when given after exposure to HPV?

a. Yes
b. No
c. Maybe
d. I don’t know
9. Can the HPV vaccine prevent infections that may lead to cervical and other HPV-related cancers?

a. Yes
b. No
c. Maybe
d. I don’t know
10. Is it necessary to test for HPV infection before administering the vaccine?

a. Yes
b. No
c. Maybe
d. I don’t know
11. Is cervical cancer one of the most common cancers affecting women in India?

a. Yes
b. No
c. Maybe
d. I don’t know
12. Is persistent infection with high-risk HPV strains the primary cause of cervical cancer?

a. Yes
b. No
c. Maybe
d. I don’t know
13. Can HPV spread through sexual contact, including vaginal, anal, and oral sex?

a. Yes
b. No
c. Maybe
d. I don’t know
14. Are males also at risk of HPV-related diseases, such as oropharyngeal and anogenital cancers?

a. Yes
b. No
c. Maybe
d. I don’t know
15. Should HPV vaccination be included in school-based immunization programs for better coverage?

a. Yes
b. No
c. Maybe
d. I don’t know
16. Can consistent condom use completely prevent HPV transmission?

a. Yes
b. No
c. Maybe
d. I don’t know
17. Does having multiple sexual partners increase the risk of acquiring HPV infection?

a. Yes
b. No
c. Maybe
d. I don’t know
18. Can HPV infection remain asymptomatic for years before causing health complications?

a. Yes
b. No
c. Maybe
d. I don’t know

Parent Questionnaire

HPV is a common virus, and some of its types can cause cervical cancer and other health problems. The HPV vaccine is designed to protect against these infections and is recommended for young people before they are exposed to the virus.

As parents, your awareness and attitudes about this vaccine are essential in ensuring your child’s health and well-being. This survey aims to assess parental knowledge, concerns, and willingness to consider HPV vaccination for their children. Your responses will help us develop better awareness programs to provide families with the information they need to make informed decisions. Your answers will remain confidential.

For each question, please choose the answer that best matches your understanding.

1. Does the HPV vaccine help protect against cervical cancer?

a. Yes
b. No
c. Maybe
d. I don’t know
2. Is the HPV vaccine recommended for children and young adults between the ages of 9 and 26?

a. Yes
b. No
c. Maybe
d. I don’t know
3. Are different types of HPV vaccines available in our country?

a. Yes
b. No
c. Maybe
d. I don’t know
4. Should women who receive the HPV vaccine still undergo cervical cancer screening later in life?

a. Yes
b. No
c. Maybe
d. I don’t know
5. Is the HPV vaccine more effective when given at a younger age, before exposure to the virus?

a. Yes
b. No
c. Maybe
d. I don’t know
6. Is it recommended to give the HPV vaccine before adolescence for better protection?

a. Yes
b. No
c. Maybe
d. I don’t know
7. Can the HPV vaccine be given to children even if they are not yet sexually active?

a. Yes
b. No
c. Maybe
d. I don’t know
8. Can individuals who are already adults still benefit from the HPV vaccine?

a. Yes
b. No
c. Maybe
d. I don’t know
9. Can the HPV vaccine prevent infections that may lead to cervical cancer and other related cancers?

a. Yes
b. No
c. Maybe
d. I don’t know
10. Is HPV infection a common cause of cervical cancer?

a. Yes
b. No
c. Maybe
d. I don’t know
11. Can HPV be spread through close personal contact, including sexual contact?

a. Yes
b. No
c. Maybe
d. I don’t know
12. Is cervical cancer one of the leading cancers affecting women in India?

a. Yes
b. No
c. Maybe
d. I don’t know
13. Are boys also at risk of HPV-related health issues, such as throat and other cancers?

a. Yes
b. No
c. Maybe
d. I don’t know
14. Should both boys and girls receive the HPV vaccine for better protection?

a. Yes
b. No
c. Maybe
d. I don’t know
15. Should HPV vaccination be included in routine school vaccination programs?

a. Yes
b. No
c. Maybe
d. I don’t know
16. Can consistent condom use completely prevent HPV infection?

a. Yes
b. No
c. Maybe
d. I don’t know
17. Does having multiple partners increase the risk of HPV infection?

a. Yes
b. No
c. Maybe
d. I don’t know
18. Can HPV infection remain unnoticed for years before causing health problems?

a. Yes
b. No
c. Maybe
d. I don’t know
19. Would you consider getting your child vaccinated against HPV if recommended by a doctor?

a. Yes
b. No
c. Maybe
20. Would more awareness about HPV and the vaccine encourage you to vaccinate your child?

a. Yes
b. No
c. Maybe

### ASSENT FORM

I ………………………………………………, do hereby understand that I have been asked to participate in a study on, ‘Knowledge, attitude and practice of Human Papilloma Virus vaccine in schools-a study in Thiruvalla municipality.’ I will be required to answer a simple questionnaire as a part of the study. I have also been told that there are no tests or expenses in this study, and I am free to refuse participation if I wish. I will fully co-operate and follow the instructions of the doctor concerned. I give my consent for the publication of the results of this study. I will not seek any reward or compensation for participating in this study.

Name of Student: ……………………………….. Signature/Thumb impression…………….

Date ………………. Name of the investigator

Consent Form - Malayalam

### INFORMED CONSENT

I Mr. / Mrs. ……………………………………………… ………………….. Father/ mother/ guardian of Solely and Voluntarily decide to participate him/her in the study ‘Knowledge, Attitude, and Practice of the Human Papillomavirus vaccine in schools- a study in the Thiruvalla municipality.’ I will give information regarding my child. I am aware that there are no additional investigations or medications planned as a part of this study. I am also aware that the information collected is confidential. I have had the opportunity to ask questions about it and any questions that I have asked have been answered to my satisfaction. I consent voluntarily to publishing the result of the research in an anonymized form and understand that my child and I have the right to withdraw from research at any time. I am ensuring that I will never ask for any benefit for participating in this research.

Name of the parent or guardian:

Signature / thumb impression:

Date:

Name of the child:

Name of the witness:

Signature of the witness:

Name of the researcher:

Signature:

Date:

Post survey student awareness class material (classes will be taken by the student with help from Faculty)

Protect Your Health: Learn About HPV and the HPV Vaccine

What is HPV?

Human Papillomavirus (HPV) is a very common virus that can infect both boys and girls. Most people get HPV at some point in their lives without even knowing it. While many HPV infections go away on their own, some types of HPV can cause serious health problems, including certain cancers. The good news is that there is a vaccine to help protect against these infections!

What is Cervical Cancer?

Cervical cancer is a type of cancer that affects the cervix, which is the lower part of the uterus in women. Some types of HPV can cause changes in the cells of the cervix, leading to cancer over time. However, with regular screening and the HPV vaccine, cervical cancer can be prevented. The HPV vaccine is one of the most effective ways to reduce the risk of cervical cancer and other HPV-related diseases.

What is the HPV Vaccine?

The HPV vaccine is a safe and effective way to protect against HPV infections that can cause cancer. It works best when given at a young age, before a person is exposed to the virus. Many countries recommend the vaccine for children and young adults, typically between the ages of 9 and 26. Getting vaccinated early provides long-lasting protection and helps prevent HPV-related diseases in the future.

Why is the HPV Vaccine Important?

The HPV vaccine helps protect you from infections that can lead to serious health problems later in life. Vaccination programs in different countries have already reduced the number of HPV infections and related diseases. Like other vaccines you receive, such as those for measles or tetanus, the HPV vaccine is an important step in staying healthy. It is most effective when given before exposure to HPV, which is why young people are encouraged to take it.

Is the HPV Vaccine Safe?

Yes! The HPV vaccine has been tested in many countries and is approved by major health organizations. Millions of people worldwide have received the vaccine, and studies have shown that it is very safe. Like any vaccine, some people may have mild side effects, such as a sore arm or slight fever, but serious side effects are extremely rare. If you have any concerns, you can talk to a doctor or a trusted adult.

Can Boys Also Get the HPV Vaccine?

Yes! The HPV vaccine is recommended for both boys and girls. While HPV is most commonly linked to cervical cancer in women, it can also cause other types of cancers in men. Vaccinating boys helps reduce the spread of HPV and protects them from HPV-related diseases.

Summary of the class

- HPV is a common virus that can cause certain types of cancer.
- The HPV vaccine is a safe and effective way to prevent HPV infections.
- The vaccine works best when given at a young age.
- Boys and girls can both benefit from the HPV vaccine.
- If you have questions, talk to your parents, teachers, or a doctor for more information.

Stay Informed, Stay Protected!

Getting vaccinated is an important step in protecting your health. If you want to learn more about HPV and the vaccine, speak to a trusted adult or healthcare professional. Together, we can prevent HPV-related diseases and build a healthier future!

Understanding HPV and the HPV Vaccine: A Guide for Teachers What is HPV?

Human Papillomavirus (HPV) is one of the most common viruses affecting both men and women. While many HPV infections go away on their own, some high-risk strains can cause serious health issues, including cervical cancer, other genital cancers, and throat cancer. HPV spreads through close contact, and most people will be exposed to it at some point in their lives. Fortunately, the HPV vaccine offers strong protection against the types of HPV that cause these diseases.

Why is HPV Vaccination Important?

Cervical cancer is one of the leading causes of cancer-related deaths among women, but it is largely preventable. HPV vaccination, when given at the recommended age, significantly reduces the risk of HPV infection and, in turn, lowers the chances of developing HPV-related cancers.

The vaccine is most effective when given before an individual is exposed to the virus, which is why it is recommended for preteens and young adolescents.

The Role of Teachers in HPV Awareness

As educators, you play a crucial role in shaping students’ understanding of health and well-being. Students and their parents may turn to you with questions about HPV and the vaccine. By providing accurate information and dispelling myths, you can help promote informed decision-making regarding vaccination. Encouraging health education programs and open discussions in schools can improve awareness and acceptance of HPV vaccination.

Is the HPV Vaccine Safe?

Yes. The HPV vaccine has been studied extensively and is endorsed by major health organizations worldwide, including the WHO and CDC. Millions of doses have been administered safely. Side effects, if any, are usually mild, such as slight soreness at the injection site or a mild fever. The benefits of vaccination far outweigh any potential risks, as it helps prevent life-threatening cancers.

Who Should Get the HPV Vaccine? The HPV vaccine is recommended for:

- Girls and boys aged 9–14 years for the best immune response and protection before exposure to the virus.
- Individuals up to 26 years of age, if they have not been vaccinated earlier.
- Some adults up to 45 years of age may also benefit from vaccination, depending on their risk factors and doctor’s recommendation.

Common Myths vs. Facts

❌ Myth: Only women need the HPV vaccine.

✔ Fact: HPV affects both men and women. Boys are also recommended to receive the vaccine to reduce HPV-related cancers and prevent the spread of the virus.

❌ Myth: HPV vaccination encourages risky behaviour.

✔ Fact: Research shows that vaccination does not influence behaviour. It is a preventive health measure, just like other vaccines.

❌ Myth: The HPV vaccine is not safe.

✔ Fact: The HPV vaccine has been thoroughly tested, and millions of doses have been safely given worldwide.

How Can Schools Support HPV Awareness?

- Host awareness sessions for students, parents, and teachers to share scientifically accurate information.
- Encourage open conversations about the importance of HPV vaccination as part of routine immunization.
- Collaborate with healthcare professionals to provide vaccination programs in schools or communities.
- Distribute educational materials to ensure students and families have access to reliable information.

Your Role in a Healthier Future

By staying informed and guiding students and parents toward credible health resources, you can help prevent HPV-related diseases and promote long-term well-being. If you have further questions, reach out to healthcare professionals or refer to trusted health organizations.

Together, we can empower students with knowledge and protect future generations from preventable diseases!

Protecting Your Child’s Health: Understanding HPV and the HPV Vaccine What is HPV?

Human Papillomavirus (HPV) is a very common virus that affects both men and women. While many infections go away on their own, some types of HPV can cause serious health problems, including cervical cancer, other cancers, and genital warts. The virus spreads through close contact, and most people will be exposed to it at some point in their lives.

Why Should My Child Get the HPV Vaccine?

The HPV vaccine is one of the best ways to protect your child from HPV-related diseases. It is most effective when given at an early age, ideally between 9 and 14 years, before exposure to the virus. By vaccinating early, you ensure that your child builds strong immunity against HPV, reducing the risk of developing HPV-related cancers in adulthood.

Is the HPV Vaccine Safe?

Yes. The HPV vaccine has been tested in many countries and is recommended by the World Health Organization (WHO) and other major health organizations. Millions of doses have been given worldwide, and studies show that the vaccine is safe and highly effective. Side effects, if any, are usually mild, such as slight pain at the injection site or a low fever. The benefits of vaccination far outweigh any risks.

Can Boys Also Get the HPV Vaccine?

Yes! The HPV vaccine is recommended for both boys and girls. While cervical cancer mainly affects women, HPV can also cause other types of cancer and health issues in men. Vaccinating boys helps protect them and also reduces the overall spread of HPV in the community.

Does My Child Need HPV Screening Before Vaccination?

No. Unlike some other vaccines, the HPV vaccine does not require any prior screening. It is designed to prevent HPV infections before they occur, making early vaccination the best strategy for protection.

Common Myths vs. Facts

❌ Myth: Only girls need the HPV vaccine.

✔ Fact: HPV affects both boys and girls. The vaccine is recommended for all children.

❌ Myth: The HPV vaccine is not necessary if my child is healthy.

✔ Fact: Even healthy children can be exposed to HPV. Vaccination is the best way to ensure long-term protection.

❌ Myth: The HPV vaccine causes serious side effects.

✔ Fact: The vaccine has been tested extensively and is proven to be safe, with only mild and temporary side effects.

How Can Parents Support HPV Awareness?

- Talk to your child about the importance of vaccination and general health protection.
- Consult your doctor to learn more about the vaccine schedule and benefits.
- Encourage vaccination as part of routine immunization for your child.
- Stay informed by referring to reliable health sources such as WHO and national health authorities.

Your Role in Protecting Your Child’s Future

As a parent, your decision to vaccinate your child against HPV can help protect them from serious diseases later in life. The HPV vaccine is a simple, safe, and effective way to ensure a healthier future for your child. If you have any questions, consult a healthcare provider for more information.

Together, we can prevent HPV-related diseases and protect our children’s health!

## Notes

### Competing Interest Statement

The authors have declared no competing interest.

### Funding Statement

This research has no funding at this time.

### Author Declarations

Institutional Ethics Committee of Pushpagiri Institute of Medical Sciences & Research Centre, Tiruvalla, Pathanamthitta, Kerala, India gave ethical approval for this work.

